# SARS-CoV-2 infection hospitalization, severity, criticality, and fatality rates

**DOI:** 10.1101/2020.11.29.20240416

**Authors:** Shaheen Seedat, Hiam Chemaitelly, Houssein Ayoub, Monia Makhoul, Ghina R. Mumtaz, Zaina Al Kanaani, Abdullatif Al Khal, Einas Al Kuwari, Adeel A. Butt, Peter Coyle, Andrew Jeremijenko, Anvar Hassan Kaleeckal, Ali Nizar Latif, Riyazuddin Mohammad Shaik, Hadi M. Yassine, Mohamed G. Al Kuwari, Hamad Eid Al Romaihi, Mohamed H. Al-Thani, Roberto Bertollini, Laith J. Abu-Raddad

## Abstract

**Background:** This study aimed to estimate the age-stratified and overall morbidity and mortality rates of the severe acute respiratory syndrome coronavirus 2 (SARS-CoV-2) infection based on an analysis of the pervasive SARS-CoV-2 epidemic in Qatar, a country with <9% of the population being ≥50 years of age.

**Methods:** Infection disease outcomes were investigated using a Bayesian approach applied to an age-structured mathematical model describing SARS-CoV-2 transmission and disease progression in the population. The model was fitted to infection and disease time-series and age-stratified data. Two separate criteria for classifying morbidity were used: one based on actual recorded hospital admission (acute-care or intensive-care-unit hospitalization) and one based on clinical presentation as per World Health Organization classification of disease severity or criticality.

**Results:** All outcomes showed very strong age dependence, with low values for those <50 years of age, but rapidly growing rates for those ≥50 years of age. The strong age dependence was particularly pronounced for infection criticality rate and infection fatality rate. Infection acute-care and intensive-care-unit bed hospitalization rates were estimated at 13.10 (95% CI: 12.82-13.24) and 1.60 (95% CI: 1.58-1.61) per 1,000 infections, respectively. Infection severity and criticality rates were estimated at 3.06 (95% CI: 3.01-3.10) and 0.68 (95% CI: 0.67-0.68) per 1,000 infections, respectively. Infection fatality rate was estimated at 1.85 (95% CI: 1.74-1.95) per 10,000 infections.

**Conclusions:** SARS-CoV-2 severity and fatality in Qatar was not high and demonstrated a very strong age dependence with <4 infections in every 1,000 being severe or critical and <2 in every 10,000 being fatal. Epidemic expansion in nations with young populations may lead to lower disease burden than previously thought.

## Introduction

The pandemic caused by the severe acute respiratory syndrome coronavirus 2 (SARS-CoV-2) infection and associated Coronavirus Disease 2019 (COVID-19) continue to be a global health challenge [1]. Aside from direct implications on morbidity and mortality [1], the pandemic led to severe economic and societal disruptions [2], a consequence of the social and physical distancing restrictions imposed to slow SARS-CoV-2 transmission in view of its severity and fatality. Had infection severity and fatality been comparable to those of seasonal influenza or other common cold coronaviruses, such restrictions may not have been deemed necessary.

One of the most affected countries by this pandemic is Qatar, a peninsula in the Arabian Gulf region with a population of 2.8 million [3, 4]. With its unique demographic and residential dwelling structure where 60% of the population are expatriate craft and manual workers (CMWs) living in large shared housing accommodations [3, 4], Qatar experienced a pervasive epidemic with >60,000 laboratory-confirmed infections per million population as of November 23, 2020 [5, 6]. The epidemic grew rapidly starting from March 2020, peaked in late May 2020, then rapidly declined in subsequent weeks, and has been in a stable low-incidence phase now for four months (Figure S1 of Supplementary Material (SM)). A series of serological studies completed by November 23, 2020 suggested that about half of the population have already been infected [5, 7-10].

With a well-resourced public healthcare structure and a centralized and standardized data-capture system for all SARS-CoV-2 testing and COVID-19 care, Qatar has one of the most extensive databases to characterize this epidemic and its toll [5]. In addition to large-scale polymerase chain reaction (PCR) and serological testing, multiple population-based PCR and serological surveys have been conducted to date [5, 7-9]. As of November 23, 2020, cumulative overall testing rates exceeded 638,000 per million population for PCR and 105,000 per million population for antibodies [11]. A comprehensive clinical characterization has also been completed for the hospitalized COVID-19 cases through individual chart reviews by trained medical personnel [11], including infection severity classification as per the World Health Organization (WHO) guidelines [12].

Given the pervasive and advanced nature of the epidemic and availability of extensive epidemiological data, Qatar provides a unique opportunity to assess the extent of SARS-CoV-2 morbidity and fatality. We aimed in this study to estimate the age-stratified and overall infection acute-care and intensive-care-unit (ICU) hospitalization rates, infection severity and criticality rates, and infection fatality rate.

## Methods

### Mathematical model and parameterization

Building on our previously developed SARS-CoV-2 models [5, 13-17], an age-structured deterministic mathematical model was constructed to describe SARS-CoV-2 transmission dynamics and disease progression in the population (Figure S2 of SM). Susceptible individuals in each age group were at risk of acquiring the infection based on their infectious contact rate per day, age-specific susceptibility to the infection, and an age-mixing matrix defining mixing between individuals in the different age groups. Following a latency period, infected individuals developed an infection that either did not require hospitalization, or that required hospitalization in an acute-care bed or in an ICU bed. Individuals admitted to an ICU bed had an additional risk for COVID-19 mortality. The model further included compartments tracking infection severity (asymptomatic/moderate/mild infection, severe infection, or critical infection as per WHO severity classification) [12]. Population movement between model compartments was described using a set of coupled nonlinear differential equations (Text S1 of SM).

The model was parameterized using best available data for SARS-CoV-2 natural history and epidemiology. Model parameters, definitions, and justifications can be found in Tables S1 and S2 of SM. The size and demographic structure of the population of Qatar were based on a population census conducted by Qatar’s Planning and Statistics Authority [4].

### Model fitting and analyses

The model was fitted to extensive time-series and age-stratified data for PCR laboratory-confirmed infections, PCR testing positivity rate, antibody testing positivity rate, PCR and serological surveys, daily hospital admissions in acute-care and ICU beds, hospital occupancy in acute-care and ICU beds, incidence of severe and critical infections as per WHO classification [12], and COVID-19 deaths (further details in Text S2 of SM). A Bayesian method, based on the incremental mixture importance sampling with shot-gun optimization [18, 19], was used to fit the model to the different data sources and to estimate the mean and 95% credible interval (CI) for each estimated parameter (Text S2 of SM). The model was coded, fitted, and analyzed using MATLAB R2019a [20].

### Outcome measures

Table 1 provides a listing of each outcome measure estimated in this study, its definition, and its interpretation. Two sets of outcome measures were generated. The first set includes *crude case rates*, such as the crude case fatality rate, calculated as the cumulative number of a disease outcome (say COVID-19 death) over the cumulative number of *documented* (that is PCR laboratory-confirmed) infections.

**Table 1.** Crude case rates and infection rates estimated in this study.

| Outcome measure | Definition | Interpretation |
| --- | --- | --- |
| **Crude case rates-data estimation** |  |  |
| 1. Crude acute-care *and* ICU bed hospitalization rate | Cumulative number of hospital admissions into acute-care or ICU beds over the cumulative number of *documented* PCR laboratory-confirmed infections | Proportion of PCR laboratory-confirmed infections that progressed to hospital admission into acute-care or ICU beds |
| 2. Crude case severity *and* criticality rate | Cumulative number of COVID-19 severe or critical infections* over the cumulative number of *documented* PCR laboratory-confirmed infections | Proportion of PCR laboratory-confirmed infections that progressed to become severe or critical |
| 3. Crude case fatality rate | Cumulative number of COVID-19 deaths over the cumulative number of *documented* PCR laboratory-confirmed infections | Proportion of PCR laboratory-confirmed infections that ended in COVID-19 death |
| **Infection rates-model estimation** |  |  |
| 1. Infection acute-care bed hospitalization rate | Cumulative number of hospital admissions into acute-care beds over the cumulative number of infections, *documented and undocumented* | Proportion of infections that progressed to acute-care bed hospital admission |
| 2. Infection ICU bed hospitalization rate | Cumulative number of hospital admissions into ICU beds over the cumulative number of infections, *documented and undocumented* | Proportion of infections that progressed to ICU bed hospital admission |
| 3. Infection severity rate | Cumulative number of COVID-19 severe infections* over the cumulative number of infections, *documented and undocumented* | Proportion of infections that progressed to become severe infections |
| 4. Infection criticality rate | Cumulative number of COVID-19 critical infections* over the cumulative number of infections, *documented and undocumented* | Proportion of infections that progressed to become critical infections |
| 5. Infection fatality rate | Cumulative number of COVID-19 deaths over the cumulative number of infections, *documented and undocumented* | Proportion of infections that ended in COVID-19 death |
| **Combined infection rates-model estimation** |  |  |
| 1. Infection acute-care *and* ICU bed hospitalization rate | Cumulative number of admissions into acute-care or ICU beds over the cumulative number of infections, *documented and undocumented* | Proportion of infections that progressed to acute-care or ICU bed hospital admission |
| 2. Infection severity *and* criticality rates | Cumulative number of COVID-19 severe or critical infections* over the cumulative number of infections, *documented and undocumented* | Proportion of infections that progressed to become severe or critical infections |

\*Per World Health Organization (WHO) infection severity classification [12].

The second set includes *infection rates*, such as the infection fatality rate, calculated as the cumulative number of a disease outcome (say COVID-19 death) over the model-estimated cumulative number of infections, *documented and undocumented*.

Two separate criteria for classifying morbidity were used: one based on actual recorded hospital admission (acute-care or ICU) and one based on clinical presentation as per WHO classification of disease severity [12]. While the two are overlapping with severe cases typically admitted to acute-care beds, and critically ill cases admitted to ICU beds, a significant fraction of mild or moderately ill cases were hospitalized out of caution. Moreover, hospitalization was used as a form of case isolation earlier in the epidemic. Of note that the health system in Qatar remained well within its threshold even at the epidemic peak towards end of May 2020.

## Results

Figure 1A shows the crude case acute-care *and* ICU bed hospitalization rate versus time. The rate was rather stable, but with a slightly declining trend, and was assessed at 113.9 acute-care *and/or* ICU hospital admissions per 1,000 laboratory-confirmed infections on November 22, 2020. As of this date, a total of 18,509 acute-care and 1,759 ICU hospital admissions had been registered.

**Figure 1.**
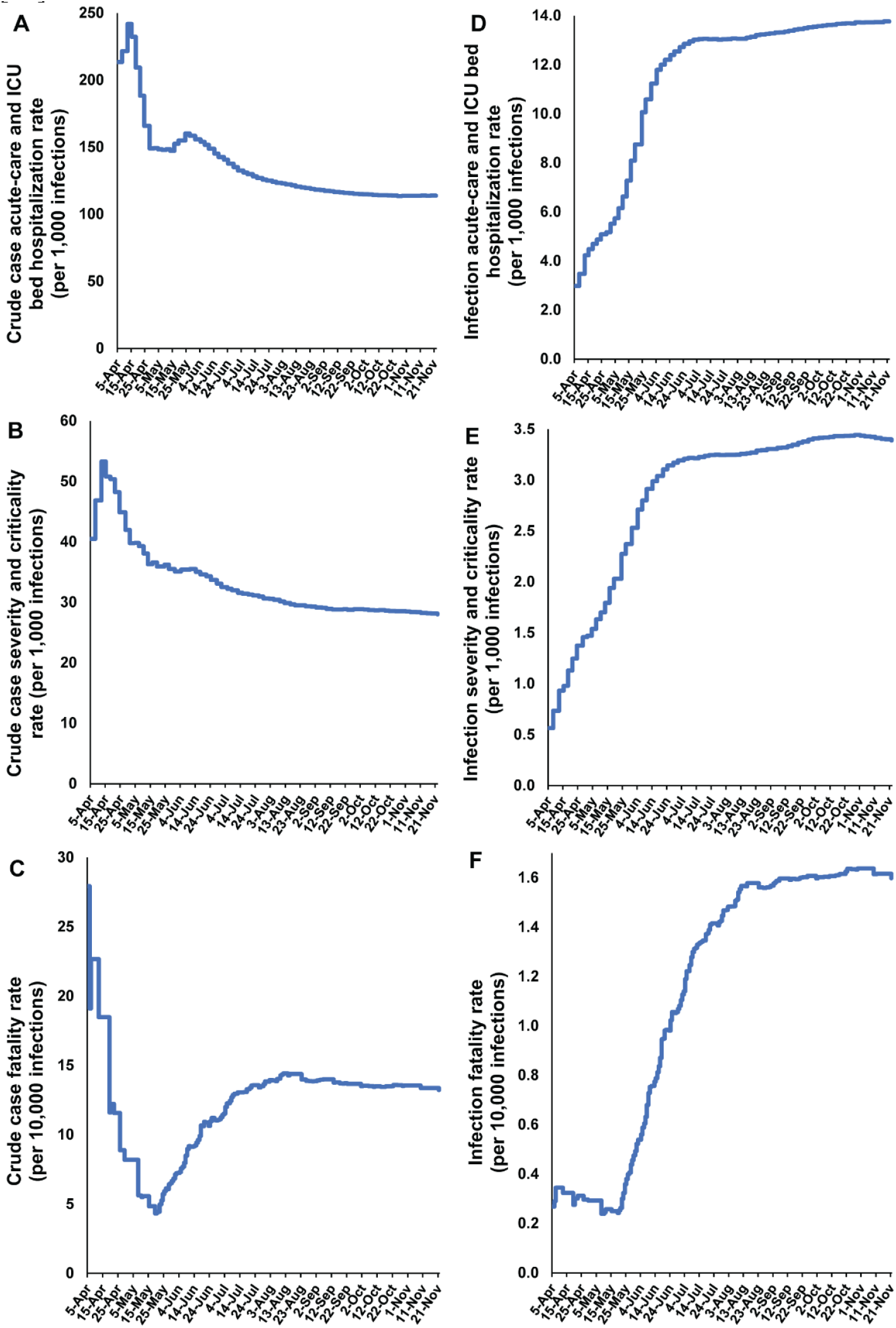
Temporal trend in A) crude case acute-care and ICU bed hospitalization rate, B) crude case severity and criticality rate, C) crude case fatality rate, D) infection acute-care and ICU bed hospitalization rate, E) infection severity and criticality rate, and F) infection fatality rate. Classification of infection severity and criticality was per WHO infection severity classification [12].

Figure 1B shows the crude case severity *and* criticality rate versus time. The rate was rather stable, but with a slightly declining trend, and was assessed at 28.0 severe *and/or* critical cases per 1,000 laboratory-confirmed infections on November 22, 2020. As of this date, a total of 4,127 severe and 863 critical infections had been registered.

Figure 1C shows the crude case fatality rate versus time. The rate increased over time, as expected with the weeks-long delay between infection and eventual COVID-19 death, but then stabilized as the epidemic entered its low but stable infection incidence phase (Figure S1 of SM). The rate was assessed at 13.2 deaths per 10,000 laboratory-confirmed infections on November 22, 2020. As of this date, a total of 235 COVID-19 deaths had been registered.

The model produced robust fits to each dataset. Table S3 of SM summarizes the goodness-of-fit. Figures S3-S7 of SM show the *age-specific* posterior distributions of the infection acute-care bed hospitalization rate (Figure S3), infection ICU bed hospitalization rate (Figure S4), infection severity rate (Figure S5), infection criticality rate (Figure S6), and infection fatality rate (Figure S7). Meanwhile, Figures S8-S9 of SM show the *overall* (total population of all age groups) infection acute-care bed hospitalization rate (Figure S8A), infection ICU bed hospitalization rate (Figure S8B), infection severity rate (Figure S8C), infection criticality rate (Figure S8D), and infection fatality rate (Figure S9).

Table 2, Figure 2, and Figure 3A show the estimated mean and 95% CI of all *age-specific infection rate* measures. All rates showed very strong age dependence. Measures increased steadily with age, with low values for those <50 years of age, but very rapidly growing rates for those ≥50 years of age. The strong age dependence was even more pronounced for infection ICU bed hospitalization rate (Figure 2B), infection criticality rate (Figure 2D), and infection fatality rate (Figure 3A).

**Table 2.** Estimated mean and 95% credible interval (CI) of the age-specific infection acute-care and ICU bed hospitalization rates, infection severity and criticality rates, and infection fatality rate. Classification of infection severity and criticality was per WHO infection severity classification [12].

| Age group (years) | Infection acute-care bed hospitalization rate (per 1,000 infections) | Infection ICU bed hospitalization rate (per 1,000 infections) | Infection severity rate (per 1,000 infections) | Infection criticality rate (per 1,000 infections) | Infection fatality rate (per 10,000 infections) |
| --- | --- | --- | --- | --- | --- |
|  | Mean (95% CI) | Mean (95% CI) | Mean (95% CI) | Mean (95% CI) | Mean (95% CI) |
| 0-9 | 8.06 (7.85-8.20) | 0.44 (0.43-0.46) | 0.07 (0.07-0.08) | 0.00 (0.00-0.00) | 0.00 (0.00-0.00) |
| 10-19 | 7.15 (6.96-7.27) | 0.39 (0.38-0.41) | 0.13 (0.13-0.13) | 0.04 (0.04-0.04) | 0.00 (0.00-0.00) |
| 20-29 | 7.19 (7.02-7.28) | 0.40 (0.39-0.41) | 0.47 (0.46-0.48) | 0.08 (0.08-0.08) | 0.12 (0.11-0.13) |
| 30-39 | 10.07 (9.88-10.18) | 0.71 (0.69-0.72) | 1.86 (1.83-1.89) | 0.21 (0.21-0.21) | 0.09 (0.09-0.10) |
| 40-49 | 16.00 (15.65-16.17) | 1.92 (1.90-1.94) | 4.05 (3.99-4.11) | 0.73 (0.72-0.73) | 0.75 (0.70-0.80) |
| 50-59 | 26.35 (25.69-26.68) | 4.67 (4.63-4.70) | 8.35 (8.22-8.48) | 2.10 (2.08-2.12) | 5.31 (4.99-5.65) |
| 60-69 | 60.46 (58.82-61.53) | 12.65 (12.54-12.75) | 25.08 (24.70-25.48) | 7.43 (7.37-7.49) | 27.68 (26.15-29.26) |
| 70-79 | 99.84 (97.23-101.65) | 38.03 (37.72-38.32) | 50.16 (49.39-50.95) | 22.45 (22.25-22.62) | 116.44 (110.43-122.83) |
| 80+ | 36.73 (35.76-37.37) | 32.94 (32.60-33.26) | 31.26 (30.78-31.76) | 23.02 (22.82-23.19) | 175.76 (168.92-183.17) |
| Overall population | 13.10 (12.82-13.24) | 1.60 (1.58-1.61) | 3.06 (3.01-3.10) | 0.68 (0.67-0.68) | 1.85 (1.74-1.95) |

**Figure 2.**
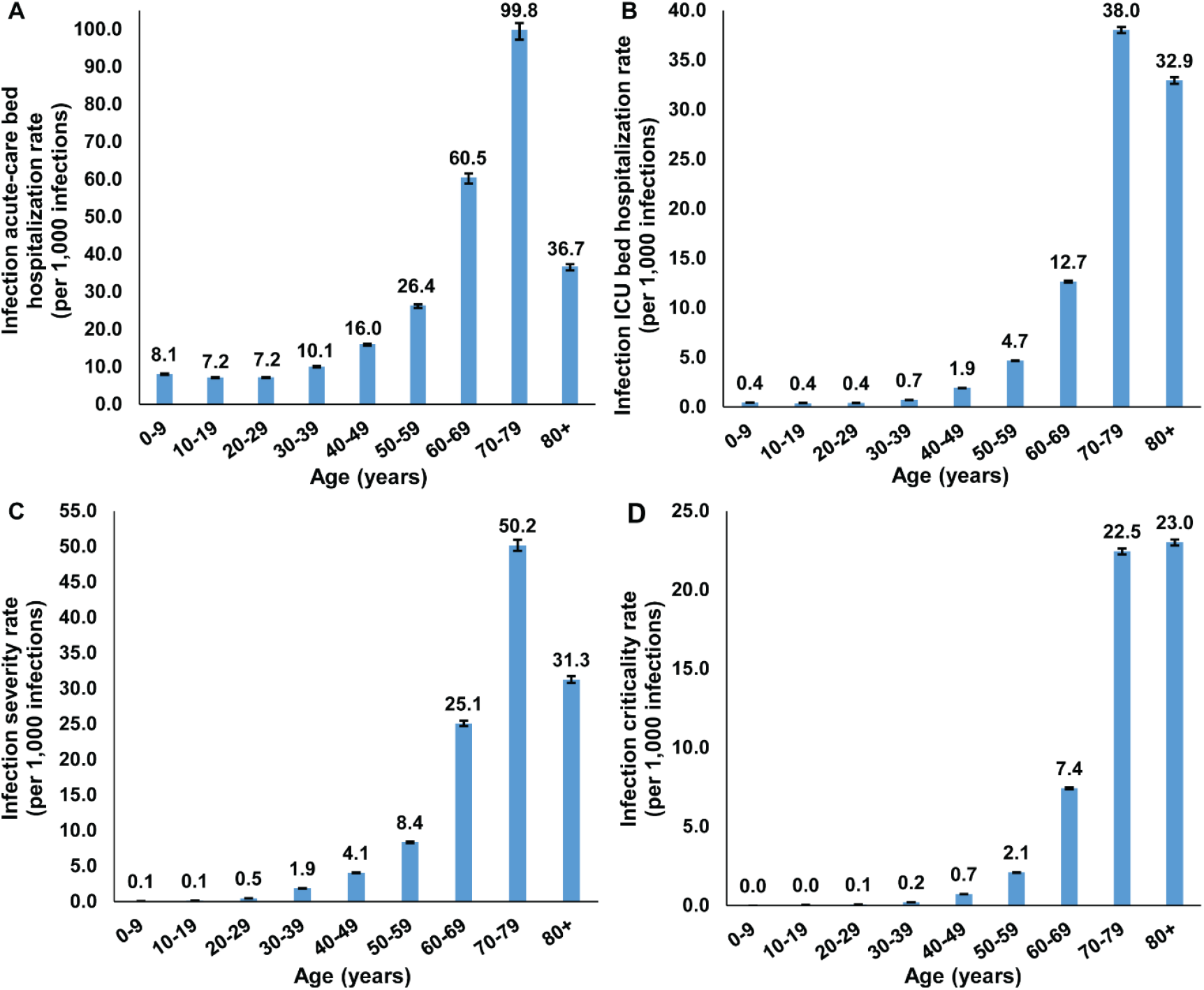
The *age-specific* A) infection acute-care bed hospitalization rate, B) infection ICU bed hospitalization rate, C) infection severity rate, and D) infection criticality rate. Classification of infection severity and criticality was per WHO severity classification [12].

**Figure 3.**
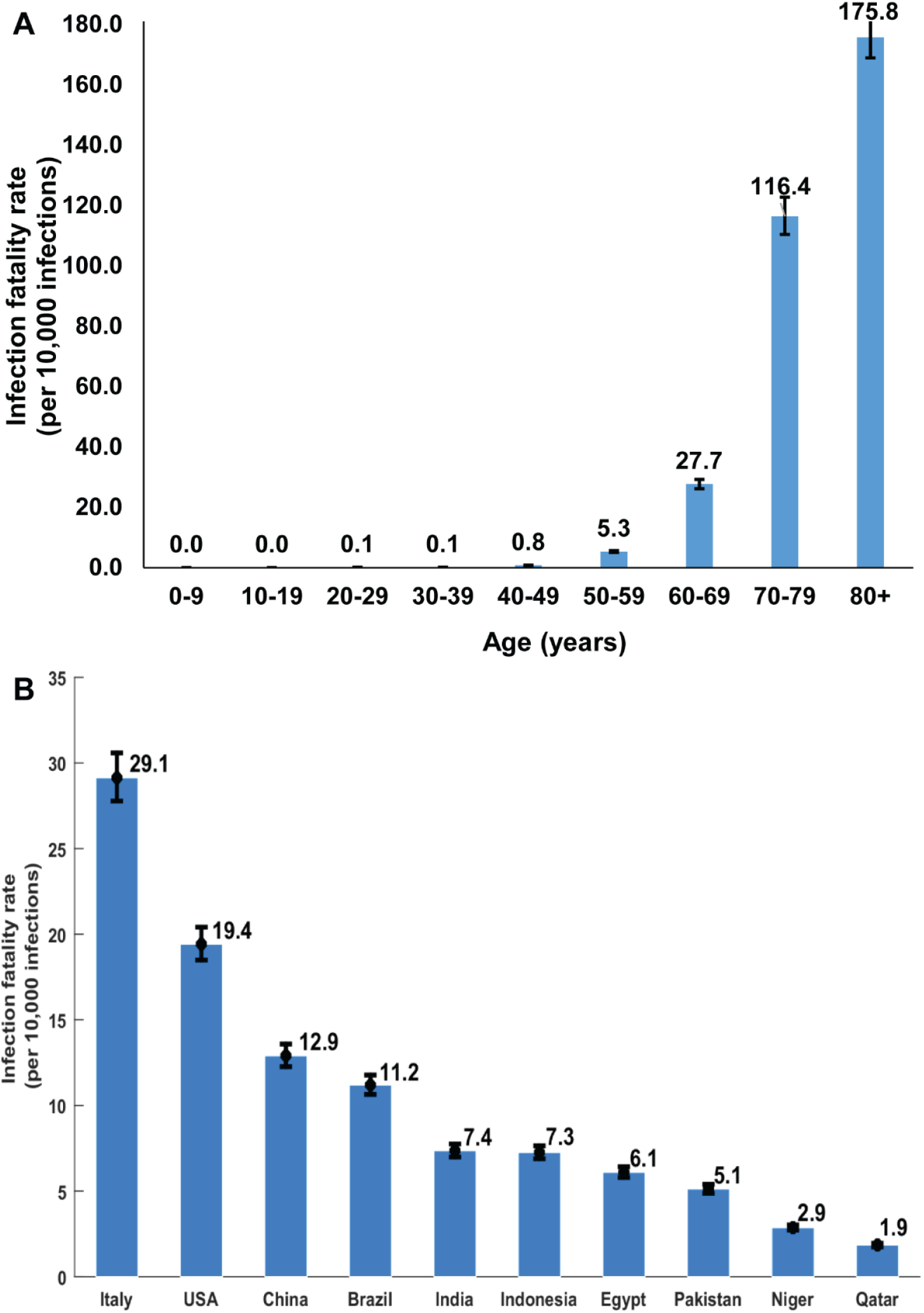
A) The *age-specific* infection fatality rate in Qatar. B) Estimated overall (total population of all age groups) infection fatality rate for several select countries that are characterized by diverse demographic structures. The estimates were generated by applying the Qatar-estimated *age-specific* infection fatality rate to the age structure of the population in each country.

The *overall* (total population of all age groups) infection acute-care bed hospitalization rate was estimated at 13.10 (95% CI: 12.82-13.24) per 1,000 infections, infection ICU bed hospitalization rate at 1.60 (95% CI: 1.58-1.61) per 1,000 infections, infection severity rate at 3.06 (95% CI: 3.01-3.10) per 1,000 infections, infection criticality rate at 0.68 (95% CI: 0.67-0.68) per 1,000 infections, and infection fatality rate at 1.85 (95% CI: 1.74-1.95) per 10,000 infections.

Figure 1D shows the infection acute-care *and* ICU bed hospitalization rate versus time. The rate increased over time, as expected with the delay between infection and hospital admission, but then stabilized (Figure 1D) as the epidemic peaked and started to decline (Figure S1 of SM). The rate was assessed at 13.8 hospital admissions per 1,000 infections on November 22, 2020. A similar pattern was found for the infection severity *and* criticality rate (Figure 1E) and infection fatality rate (Figure 1F), which were assessed at 3.4 cases per 1,000 infections and 1.6 deaths per 10,000 infections, respectively, on November 22, 2020.

The study generated other relevant estimates. The diagnosis (detection) rate as of November 22, 2020, that is the proportion of infections that were documented (with PCR laboratory-confirmed diagnosis) out of all infections that were estimated to have occurred, was assessed at 12.1% (95% CI: 12.0-12.1%). The mean duration of acute-care hospitalization was estimated at 8.73 (95% CI: 8.62-8.83) days and the mean duration of ICU hospitalization was estimated at 12.30 (95% CI: 12.18-12.41) days.

## Discussion

The striking finding of this study is that SARS-CoV-2 morbidity and mortality demonstrated a very strong age dependence. Infection severity, criticality, and fatality increased very rapidly with age. This was particularly the case for infection criticality and fatality which were limited for those <50 years of age but increased very rapidly for those ≥50 years of age (Figures 2-3). This strong age dependence combined with the lower infection exposure in those ≥60 years of age (Figure S10 of SM) and the small proportion of the population ≥50 years of age (9%) and ≥60 years of age (2%), all contributed together to a low morbidity and mortality in Qatar. Out of every 1,000 infections, only 3.7 infections were destined to be severe or critical, and out of every 10,000 infections, only 1.9 infections were destined to end in COVID-19 death (Table 2).

Notably, both SARS-CoV-2 morbidity and mortality in Qatar were not much higher than those typically seen in a seasonal influenza epidemic in the United States [21, 22]. This fact, however, needs to be interpreted in context. With the young age structure of the population in Qatar, a seasonal influenza epidemic in this country has a much lower severity than that in the United States. Typically, only a handful of influenza-related deaths are reported every year in Qatar [23].

Other lines of evidence support these findings. In a survey of ten CMW communities in Qatar, only five severe infections and one critical infection ever occurred in 3,233 persons with confirmed infection (antibody and/or PCR positive result), that is an infection severity *and* criticality rate of 2.5 (95% CI: 1.1-4.9) per 1,000 infections [7]. In another nationwide survey of the CMW population, only seven severe infections and one critical infection ever occurred in 1,590 persons with antibody and/or PCR positive result, an infection severity *and* criticality rate of 5.0 (95% CI: 2.2-9.9) per 1,000 infections [8]. Both of these estimates are in agreement with the present study estimate of 3.7 (95% CI: 3.7-3.8) per 1,000 infections.

These figures, however, are substantially lower than those estimated elsewhere, often using early epidemic data [24-29]. The fact that the early phases of the epidemic in Europe and the United States heavily affected nursing facilities and care homes of the elderly may have biased the estimates to higher values. It is also possible that our estimates are lower because of the robust accounting of the large pool of undocumented infections in the present study, thanks to the series of serological surveys conducted in Qatar [5, 7-10]. These surveys provided some of the key input data for this modeling study. For instance, the nationwide survey of the CMW population found that only 9.3% (95% CI: 7.9-11.0%) of those antibody positive had a prior documented PCR laboratory-confirmed infection [8]. This is in agreement with the diagnosis (detection) rate estimated here at 12.1% (95% CI: 12.0-12.1%), as well as growing evidence from other countries indicating that only one in every 10 infections was ever diagnosed [24, 30-33]. The totality of evidence on the Qatar epidemic also indicates that most infections were asymptomatic or minimally mild to be diagnosed [5-9, 34-36]. For instance, a national SARS-CoV-2 PCR survey found that 58.5% of those testing PCR positive reported no symptoms within the preceding two weeks of the survey [5].

In light of these findings, it is evident that the strong age dependence of SARS-CoV-2 morbidity and mortality is a principal contributor to the low morbidity and mortality seen in Qatar compared to elsewhere. The impact of this strong age dependence is illustrated in Figure 3B where the age-specific infection fatality rate of Table 2 has been applied to the age structure of other national populations. The infection fatality rate in Italy was found to be 10-fold higher than that in Qatar, *only because* of the population’s different age structure. These findings indicate that the infection morbidity and mortality may vary immensely across countries, and will be substantially lower in countries with younger demographic structures, as suggested earlier [13].

While age appears to be the principal factor, other factors could have also contributed to explaining the low morbidity and mortality in Qatar. Growing evidence indicates T cell and antibody reactivity against SARS-CoV-2 in unexposed individuals [37-40], that probably reflects development of T cell and antibody immune memory to circulating ‘common cold’ coronaviruses, which may have in turn led to lower morbidity and mortality [37-40]. The shared-housing dwelling structure in Qatar that contributed to the large SARS-CoV-2 epidemic [5] may have, along with the frequent international travel of Qatar’s expatriate population, also contributed to higher levels of exposure to other common cold coronaviruses [41, 42], thereby inducing high levels of broadly cross-reactive T cell and antibody responses. Such pre-existing immune reactivity [37-40] may have thus resulted in lower levels of SARS-CoV-2 morbidity and mortality in the population of Qatar.

The resourced healthcare system, which was well below its threshold even at the epidemic peak, may have also contributed to the low observed morbidity and mortality. Emphasis on proactive early treatment, in addition to a cautious approach for SARS-CoV-2 case management, may have limited the number of people who went on to require hospitalization, or to develop severe or critical disease.

Limitations may have affected this study. We estimated rates of infection morbidity and mortality, not accounting for potential differences by sex, comorbidities, or other factors. Model estimations are contingent on the validity and generalizability of input data. Available input data were most complete at the national level, and although there could be subpopulation differences in the highly diverse population of Qatar, these could not be factored in the model given insufficient data at the subpopulation level. Despite these limitations, our model, tailored to the complexity of the epidemic in Qatar, was able to reproduce the observed epidemic trends, and provided profound insights about healthcare needs and infection morbidity and mortality.

In conclusion, SARS-CoV-2 morbidity and mortality demonstrate a strikingly strong age dependence. With its young population structure, both morbidity and mortality were low in Qatar, and not much higher than those typically seen in a seasonal influenza epidemic in the United States [21, 22]. Out of every 1,000 infections, only 3.7 infections were severe or critical, and out of every 10,000 infections, only 1.9 infections were fatal. However, these rates would have been much higher if the population of Qatar had a similar demographic structure to that found in Europe or the United States. These findings suggest that SARS-CoV-2 morbidity and mortality may vary immensely by country or region, and that the pandemic expansion in nations with young populations may lead to much milder disease burden than currently believed.

## Data Availability

All data are available within the manuscript and its supplementary material.

## Authors’ contributions

LJA conceived and designed the study and led the mathematical modeling and statistical analyses and writing of the manuscript. SS co-constructed and parameterized the mathematical model, conducted the mathematical modeling analyses, and co-wrote the first draft of the manuscript. HC contributed to the parameterization of the mathematical model, conducted the statistical analyses, and co-wrote the first draft of the manuscript. HHA co-constructed and parameterized the mathematical model. All authors contributed to data collection and acquisition, database development, discussion and interpretation of the results, and to the writing of the manuscript. All authors have read and approved the final manuscript.

## Conflicts of interests

We declare no competing interests.

## Acknowledgement

We thank Her Excellency Dr. Hanan Al Kuwari, Minister of Public Health, for her vision, guidance, leadership, and support. We also thank Dr. Saad Al Kaabi, Chair of the System Wide Incident Command and Control (SWICC) Committee for the COVID-19 national healthcare response, for his leadership, analytical insights, and for his instrumental role in enacting data information systems that made these studies possible. We further extend our appreciation to the SWICC Committee and the Scientific Reference and Research Taskforce (SRRT) members for their informative input, scientific technical advice, and enriching discussions. We also thank Dr. Mariam Abdulmalik, CEO of the Primary Health Care Corporation and the Chairperson of the Tactical Community Command Group on COVID-19, as well as members of this committee, for providing support to the teams that worked on the field surveillance. We further thank Dr. Nahla Afifi, Director of Qatar Biobank (QBB), Ms. Tasneem Al-Hamad, Ms. Eiman Al-Khayat and the rest of the QBB team for their unwavering support in retrieving and analyzing samples and in compiling and generating databases for COVID-19 infection, as well as Dr. Asma Al-Thani, Chairperson of the Qatar Genome Programme Committee and Board Vice Chairperson of QBB, for her leadership of this effort. We also acknowledge the dedicated efforts of the Clinical Coding Team and the COVID-19 Mortality Review Team, both at Hamad Medical Corporation, and the Surveillance Team at the Ministry of Public Health.

## Funding

The authors are grateful for support provided by the Biomedical Research Program and the Biostatistics, Epidemiology, and Biomathematics Research Core, both at Weill Cornell Medicine-Qatar, the Ministry of Public Health, and Hamad Medical Corporation. The modeling infrastructure was made possible by NPRP grant number 9-040-3-008 from the Qatar National Research Fund (a member of Qatar Foundation). GM acknowledges support by UK Research and Innovation as part of the Global Challenges Research Fund, grant number ES/P010873/1. The statements made herein are solely the responsibility of the authors.

## Supplementary Material

**Figure S1.**
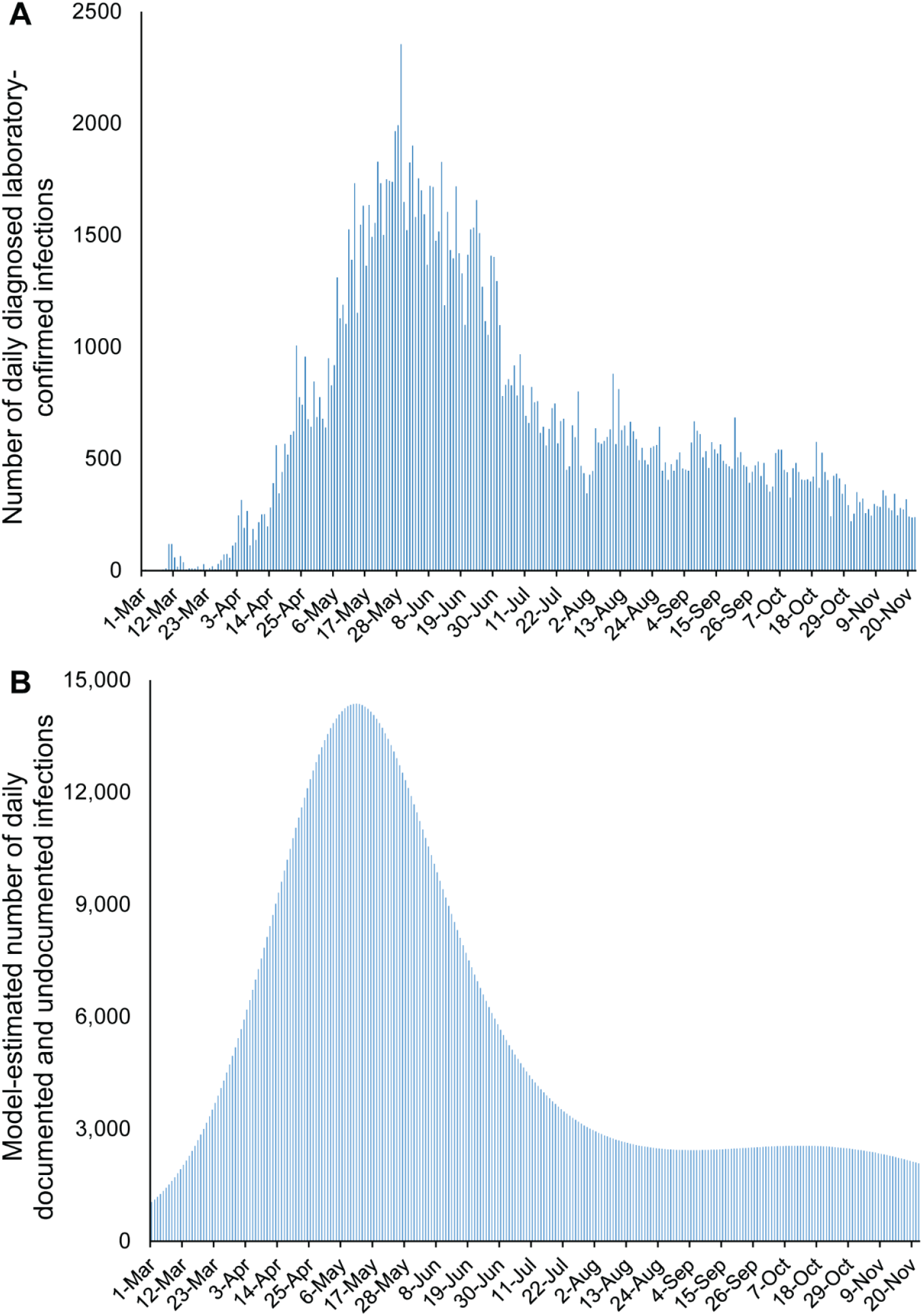
Time course of the SARS-CoV-2 epidemic in Qatar. Daily incidence of (A) documented laboratory-confirmed infections and (B) documented and undocumented infections estimated by the model.

### Text S1. Mathematical model structure and description

We constructed an age-structured deterministic mathematical model to describe the severe acute respiratory syndrome coronavirus 2 (SARS-CoV-2) transmission dynamics and disease progression in the population of Qatar (Figure S1). The model stratified the population into compartments according to age group (0-9, 10-19, 20-29,…, ≥80 years), infection stage (does not require hospitalization, requires acute-care hospitalization, requires intensive care unit (ICU) hospitalization), and hospitalization stage (acute-care hospitalization, ICU hospitalization). The model also includes five tracking compartments to track infection severity (asymptomatic/moderate/mild infection, severe infection, severe disease, critical infection, critical disease per World Health Organization (WHO) severity classification [1]). The model extends our earlier mathematical models developed to characterize SARS-CoV-2 epidemics [2-5].

Epidemic dynamics were described using age-specific sets of coupled nonlinear differential equations. Each age group, *a*, denoted a ten-year age band apart from the last category which grouped together all individuals ≥80 years of age. Qatar’s population size and demographic structure were based on findings of “The Simplified Census of Population, Housing, and Establishments” conducted by Qatar’s Planning and Statistics Authority [6]. Life expectancy was obtained from the United Nations World Population Prospects database [7].

**Figure S2.**
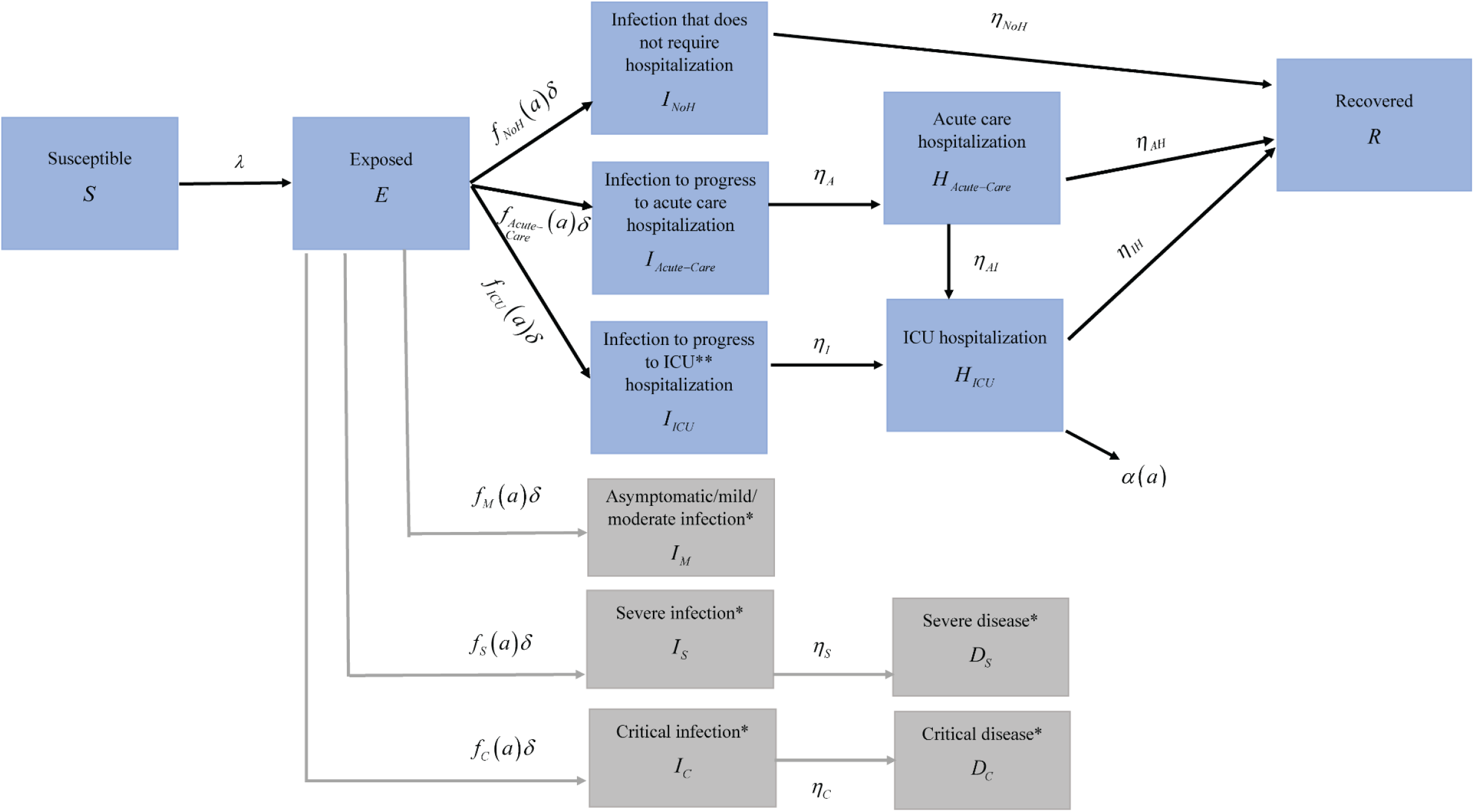
Schematic diagram describing the basic structure of the SARS-CoV-2 mathematical model and its associated tracking compartments. ^*\**^*Per World Health Organization (WHO) infection severity classification [1]* ^*\*\**^*ICU: intensive care unit*

The following equations were used to describe the transmission dynamics in the total population:

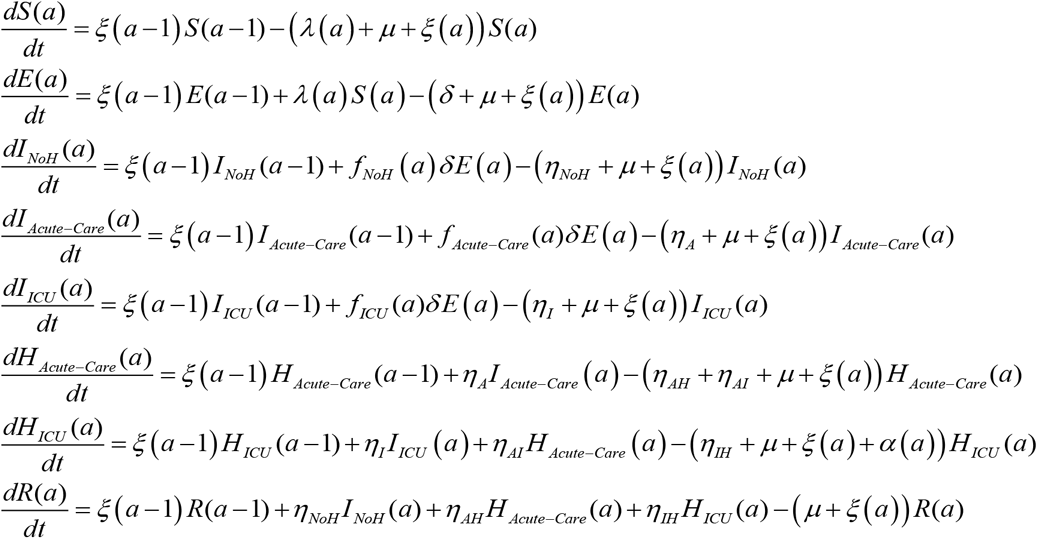

The following equations were used to track the individuals who have asymptomatic/moderate/mild infection, severe infection, severe disease, critical infection, and critical disease per WHO infection severity classification [1]:

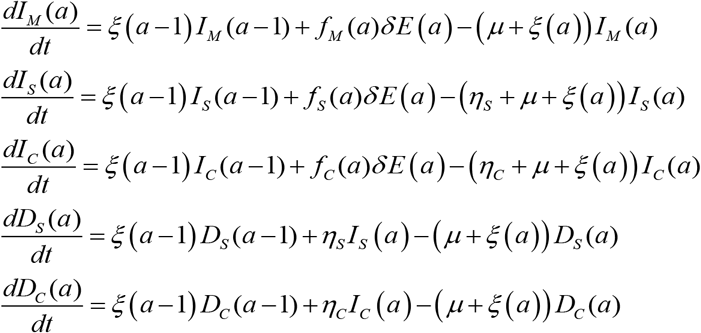

The definitions of population variables and symbols used in the equations are listed in Table S1.

**Table S1.** Definitions of population variables and symbols used in the model.

| Symbol | Definition |
| --- | --- |
| $S(a)$ | Susceptible population |
| $E(a)$ | Latently infected population |
| $I_{NoH}(a)$ | Population that is infectious but with an infection that will not require hospitalization |
| $I_{Acute-Care}(a)$ | Population that is infectious and that will progress to acute-care hospitalization |
| $I_{ICU}(a)$ | Population that is infectious and that will progress to ICU hospitalization |
| $H_{Acute-Care}(a)$ | Population with hospitalization in acute-care beds |
| $H_{ICU}(a)$ | Population with hospitalization in ICU beds |
| $R(a)$ | Recovered population |
| $I_M(a)$ | Population with an asymptomatic/moderate/mild infection per WHO infection severity classification [1] |
| $I_S(a)$ | Population with an infection that will progress to severe disease per WHO infection severity classification [1] |
| $I_C(a)$ | Population with an infection that will progress to critical disease per WHO infection severity classification [1] |
| $D_S(a)$ | Population with severe disease per WHO infection severity classification [1] |
| $D_C(a)$ | Population with critical disease per WHO infection severity classification [1] |
| $n_{age}$ | Number of age groups |
| $\xi(a)$ | Transition rate from one age group to the next age group. Here $\xi(0) = \xi(n_{age}) = 0$ |
| $\sigma(a)$ | Susceptibility profile to the infection in each age group |
| $1/\delta$ | Duration of latent infection before onset of infectiousness |
| $\beta$ | Average rate of infectious contacts |
| $1/\eta_{NoH} = 1/\eta_A = 1/\eta_I$ | Duration of infectiousness |
| $1/\eta_{AH}$ | Duration of acute-care hospitalization following hospital admission and prior to recovery |
| $1/\eta_{IH}$ | Duration of ICU hospitalization following hospital admission and prior to recovery |
| $1/\eta_{AI}$ | Duration of acute-care hospitalization prior to admission to ICU for those infected individuals that are transferred from acute-care to ICU |
| $1/\eta_S$ | Duration of severe infection prior to onset of severe disease |
| $1/\eta_C$ | Duration of critical infection prior to onset of critical disease |
| $\mu$ | Natural death rate |
| $\alpha(a)$ | Disease mortality rate in each age group |
| $f_{NoH}(a)$ | Proportion of infections that will progress to be infections that will not require hospitalization |
| $f_{Acute-Care}(a)$ | Proportion of infections that will progress to be infections that require hospitalization in acute-care beds |
| $f_{ICU}(a)$ | Proportion of infections that will progress to be infections that require hospitalization in ICU beds |
| $f_M(a)$ | Proportion of infections that are asymptomatic/moderate/mild per WHO infection severity classification [1] |
| $f_S(a)$ | Proportion of infections that will progress to be severe infections per WHO infection severity classification [1] |
| $f_C(a)$ | Proportion of infections that will progress to be critical infections per WHO infection severity classification [1] |

The force of infection (hazard rate of infection) experienced by each susceptible population, *S* (*a*), is given by

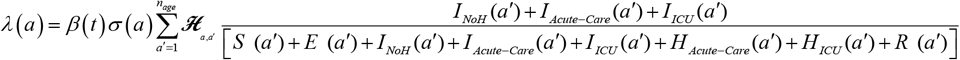

Here, *β* (*t*) is the time-dependent average rate of infectious contacts and *σ* (*a*) is the susceptibility profile to the infection in each age group *a*. To account for temporal variation in the basic reproduction number, we incorporated a temporal variation in *β* that was parameterized through a combination of Woods-Saxon and Logistic functions.

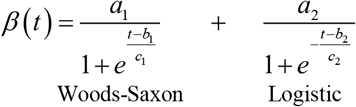

This function was mathematically designed to describe and characterize the time evolution of the level of risk of exposure before and after easing of restrictions. It was informed by our knowledge of SARS-CoV-2 epidemiology in Qatar [3], and it provided a robust fit to the data. Here *a*_1_, *a*_2_, *b*_1_, *b*_2_, *c*_1_, and *c*_2_ are fitting parameters.

The probability that an individual in the *a* age group will mix with an individual in the *a*′ age ***ℋ***_*a,a*′,_ group is determined by an age-mixing matrix,, given by

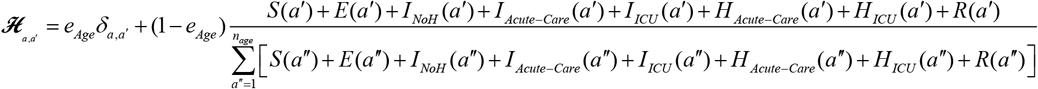

Here, *δ*_*a,a*′_ is the identity matrix and *e*_*Age*_ ∈[0,1] measures the degree of assortativeness in the age mixing. At the extreme *e*_*Age*_ = 0, the mixing is fully proportional, while at the other extreme, *e*_*Age*_ = 1, the mixing is fully assortative, that is individuals mix only with members in their own age group.

### Text S2. Parameter values, data input, and model fitting

Model input parameters were based on best available empirical data for SARS-CoV-2 natural history and epidemiology. Model input parameter values and supporting evidence are listed in Table S2.

**Table S2.** Model input parameters.

| Parameter | Symbol | Value | Justification |
| --- | --- | --- | --- |
| Duration of latent infection | $1/\delta$ | 3.69 days | Based on existing estimate [8] and based on a median incubation period of 5.1 days [9] adjusted by observed viral load among infected persons [10] and reported transmission before onset of symptoms [11] |
| Duration of infectiousness | $1/\eta_{NoH}$ ;<br>$1/\eta_A$ ;<br>$1/\eta_I$ | 3.48 days | Based on existing estimate [8] and based on observed time to recovery among persons with mild infection [8, 12] and observed viral load in infected persons [10, 11, 13] |
| Life expectancy in Qatar | $1/\mu$ | 80.7 years | United Nations World Population Prospects database [7] |

The model was fitted to extensive sources of data thanks to the centralized and standardized databases of SARS-CoV-2 testing, infection, COVID-19 disease, hospitalization, and severity as well as to findings of ongoing epidemiologic studies in Qatar. Data included: 1) time-series of number of PCR laboratory-confirmed SARS-CoV-2 infections, 2) distribution of PCR laboratory-confirmed infections by age group, 3) time-series of SARS-CoV-2 PCR testing positivity rate, 4) fraction of PCR laboratory-confirmed SARS-CoV-2 infected persons aged >60 years old, 5) a series of PCR and serological surveys, 6) age-distribution of SARS-CoV-2 antibody positivity, 7) time-series of new/daily hospital admissions in acute-care beds and in ICU beds, 8) age distribution of hospital admissions in acute-care beds and in ICU beds, 9) fraction of individuals admitted to ICU beds from acute-care beds, 10) time-series of current hospital occupancy in acute-care beds and in ICU beds, 11) time-series of new/daily severe infections and critical infections per WHO infection severity classification [1], 12) age distribution of severe and critical infections per WHO infection severity classification [1], 13) time series of COVID-19 deaths, and 14) age distribution of COVID-19 deaths.

A Bayesian method, based on incremental mixture importance sampling with shotgun optimization [14, 15], was used to conduct the model fitting and generate posteriors and credible sets. In the first stage (initialization), the importance sampling distribution was initialized by drawing a large number of points from the prior distribution and weights were attached to each point using their respective log-likelihood.

In the second stage (shot-gun optimization), multiple sequential optimizations were carried out. In each of these, the point of the importance sampling distribution with the maximum weight was computed, and a new sample set was drawn from a Gaussian distribution centered at this point. The local maximum of this sample set (point with largest log-likelihood) was then identified. The importance sampling distribution was then updated to exclude the points that are farthest from this local maximum in terms of Mahalanobis distance. A new sample set was then drawn from a Gaussian distribution centered at the current local maximum and was added to the importance sampling distribution. At the end of the shot-gun optimization stage, a candidate importance sampling distribution was obtained.

In the third stage (importance sampling), weights were attached to each point in the importance sampling distribution and sampling was iterated until convergence in the distribution of the importance sampling distribution. In the fourth stage (resampling), points were resampled from the importance sampling distribution to construct the posterior distribution of each estimated parameter.

**Table S3.** Goodness-of-fit in terms of likelihood of the model fitting each dataset.

| **Empirical dataset** | **Model fitting likelihood** |
| --- | --- |
| Time-series of number of PCR laboratory-confirmed SARS-CoV-2 infections | 80.52% |
| Distribution of PCR laboratory-confirmed infections by age group | 82.34% |
| Time-series of SARS-CoV-2 PCR testing positivity rate | 81.81% |
| Fraction of PCR laboratory-confirmed SARS-CoV-2 infections aged >60 years | 98.10% |
| Series of PCR surveys | 89.31% |
| Age-distribution of SARS-CoV-2 antibody positivity | 99.17% |
| Time-series of new/daily hospital admissions in acute-care and in ICU beds | 98.87% |
| Age distribution of hospital admissions in acute-care and in ICU beds | 99.95% |
| Fraction of ICU admissions that are being transferred from acute-care beds | 99.99% |
| Time-series of current hospital occupancy in acute-care and in ICU beds | 93.53% |
| Time-series of new/daily severe infections and critical infections* | 99.43% |
| Age distribution of severe and critical infections* | 99.96% |
| Age distribution of COVID-19 deaths | 99.98% |

\*Per World Health Organization (WHO) infection severity classification [1]

**Figure S3.**
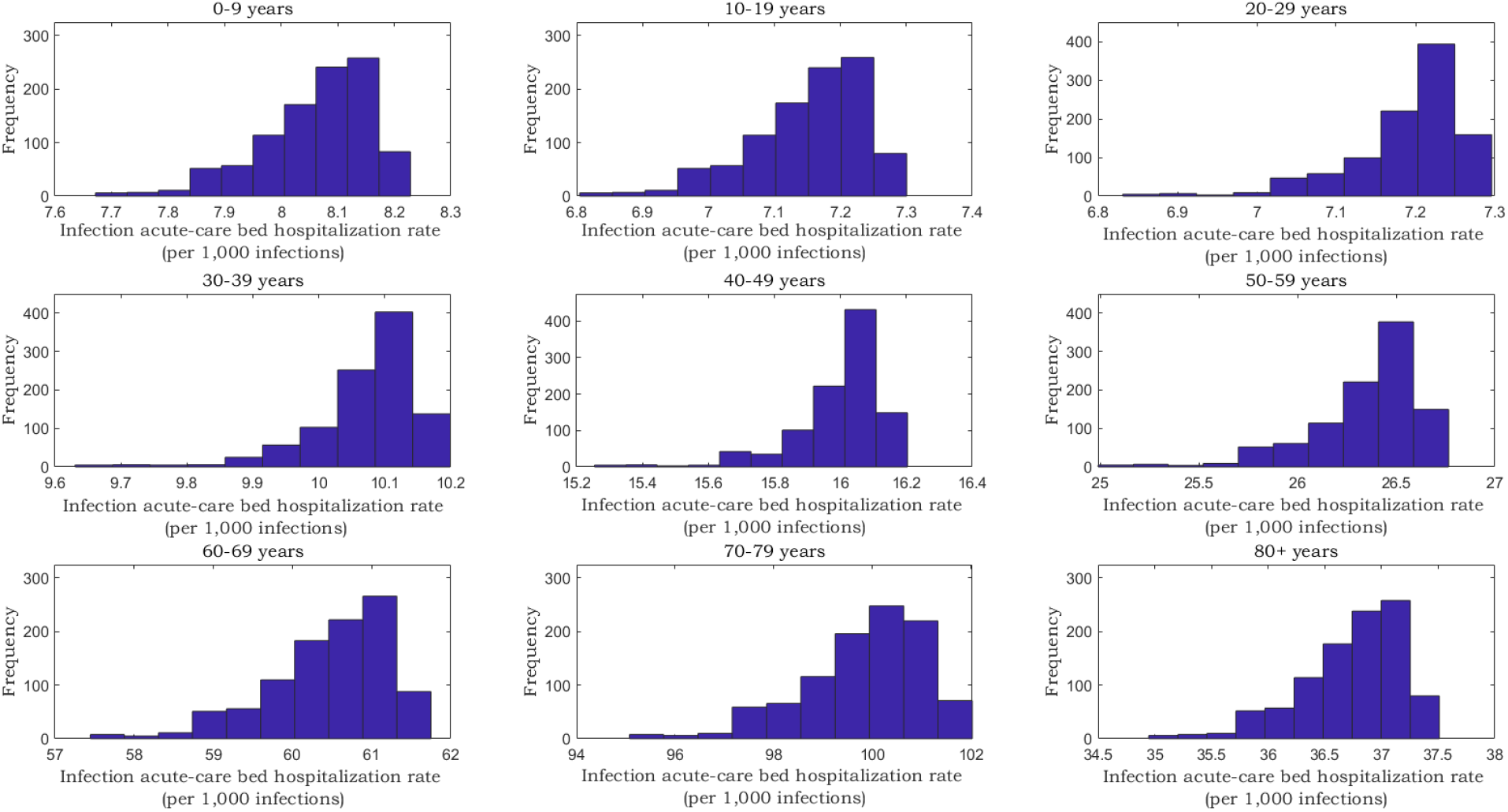
Posterior distribution of the age-specific infection acute-care bed hospitalization rate.

**Figure S4.**
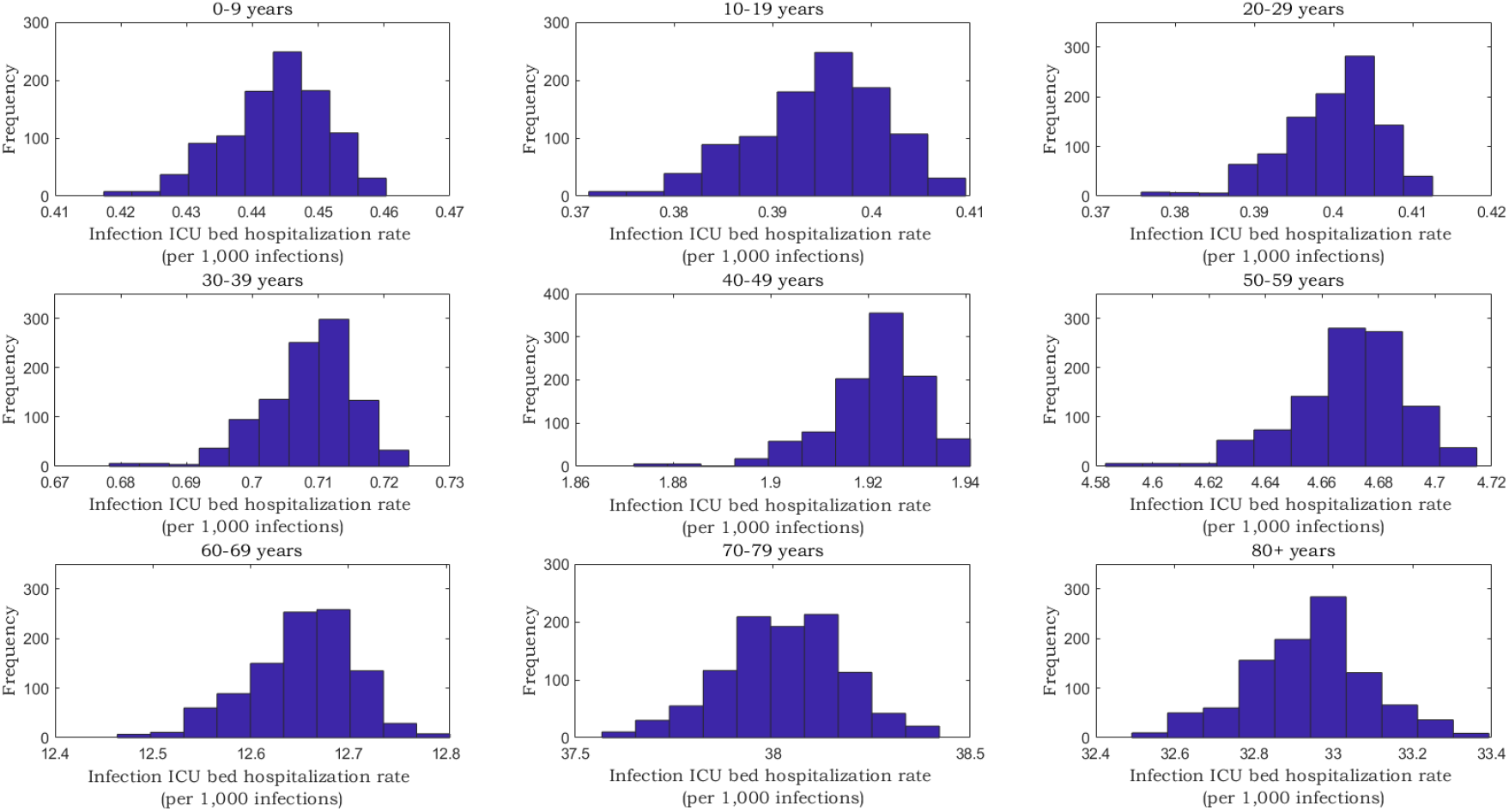
Posterior distribution of the age-specific infection ICU bed hospitalization rate.

**Figure S5.**
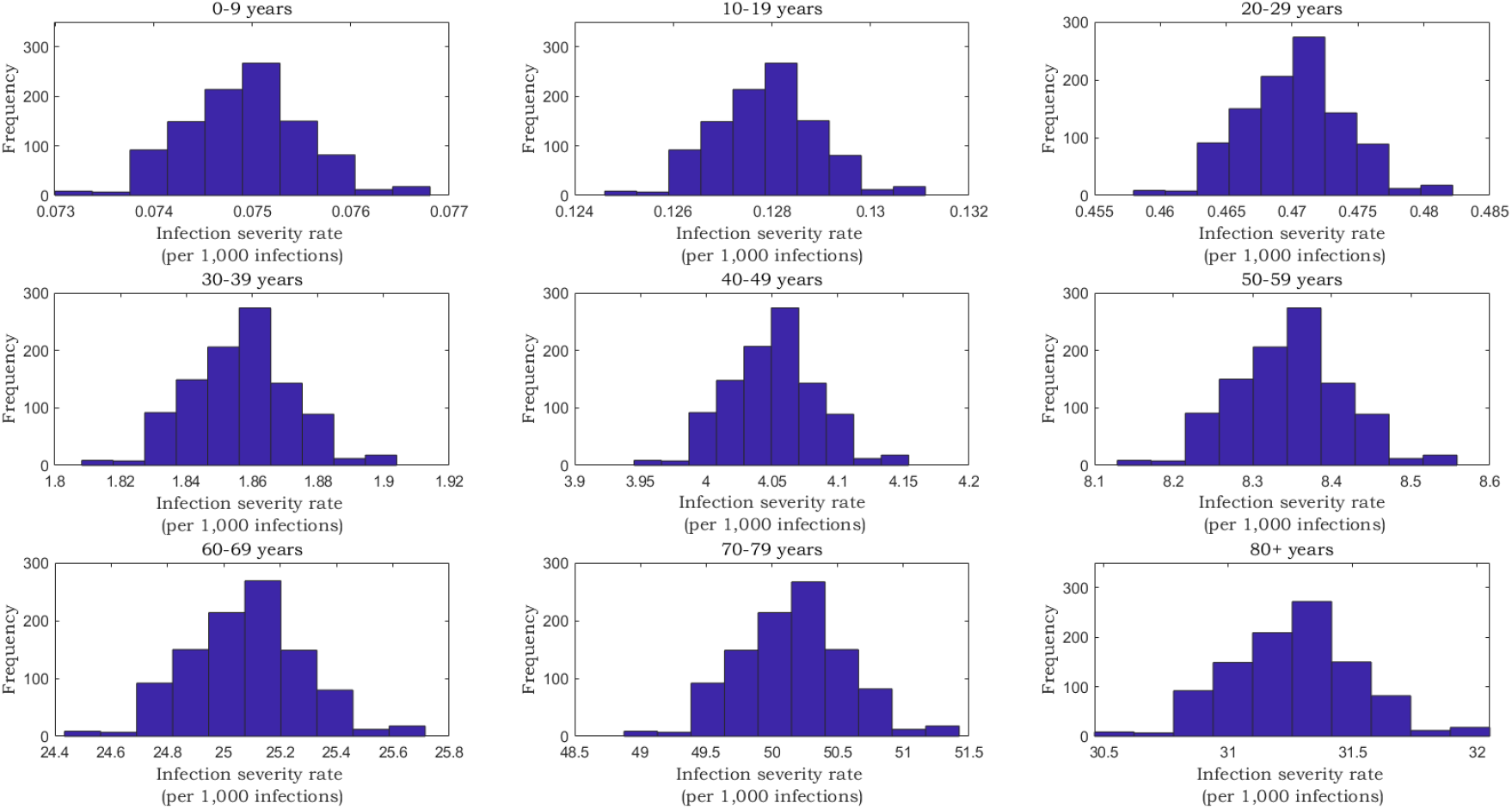
Posterior distribution of the age-specific infection severity rate. Classification of infection severity was per WHO severity classification [1].

**Figure S6.**
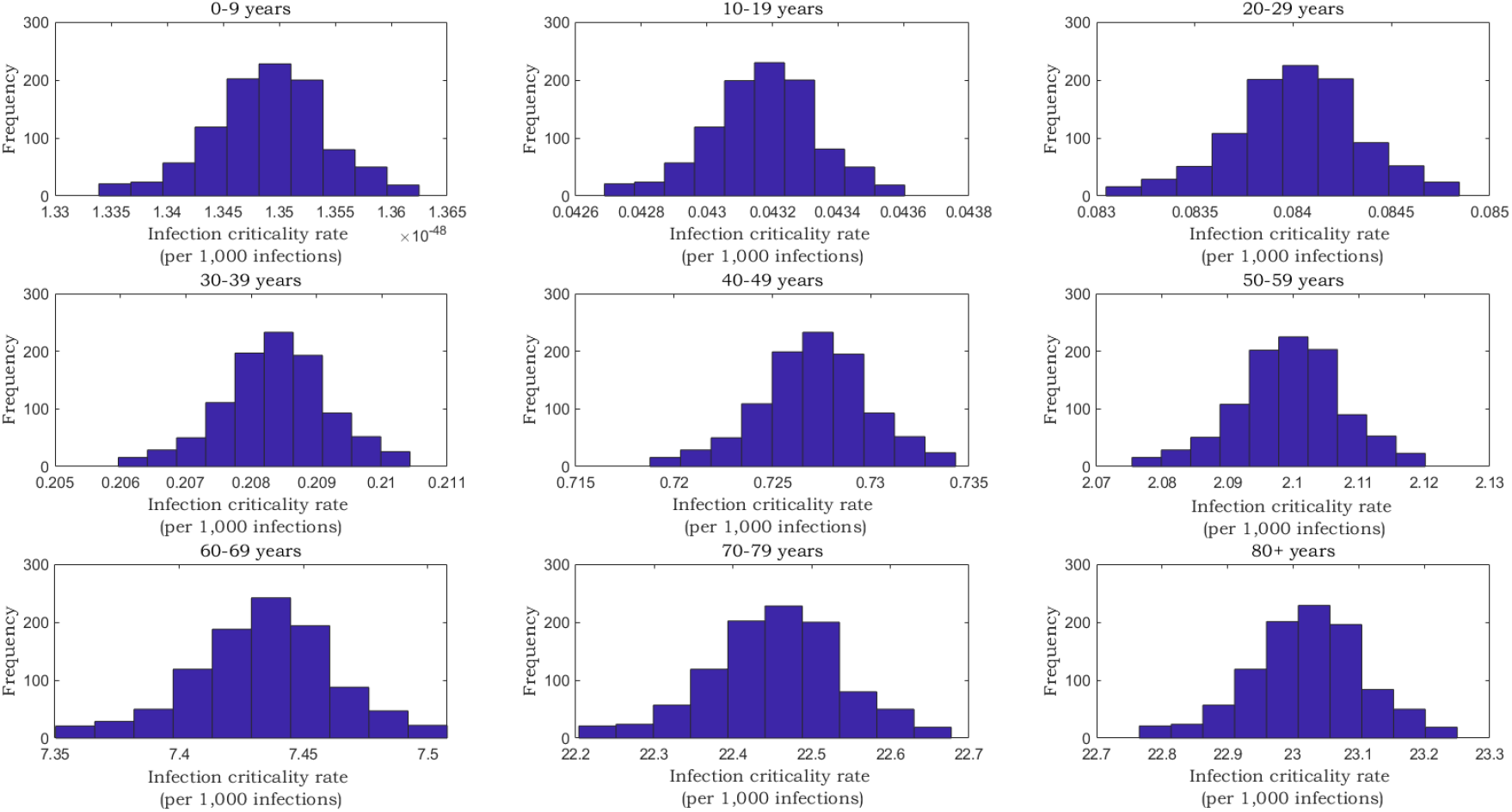
Posterior distribution of the age-specific infection criticality rate. Classification of infection criticality was per WHO severity classification [1].

**Figure S7.**
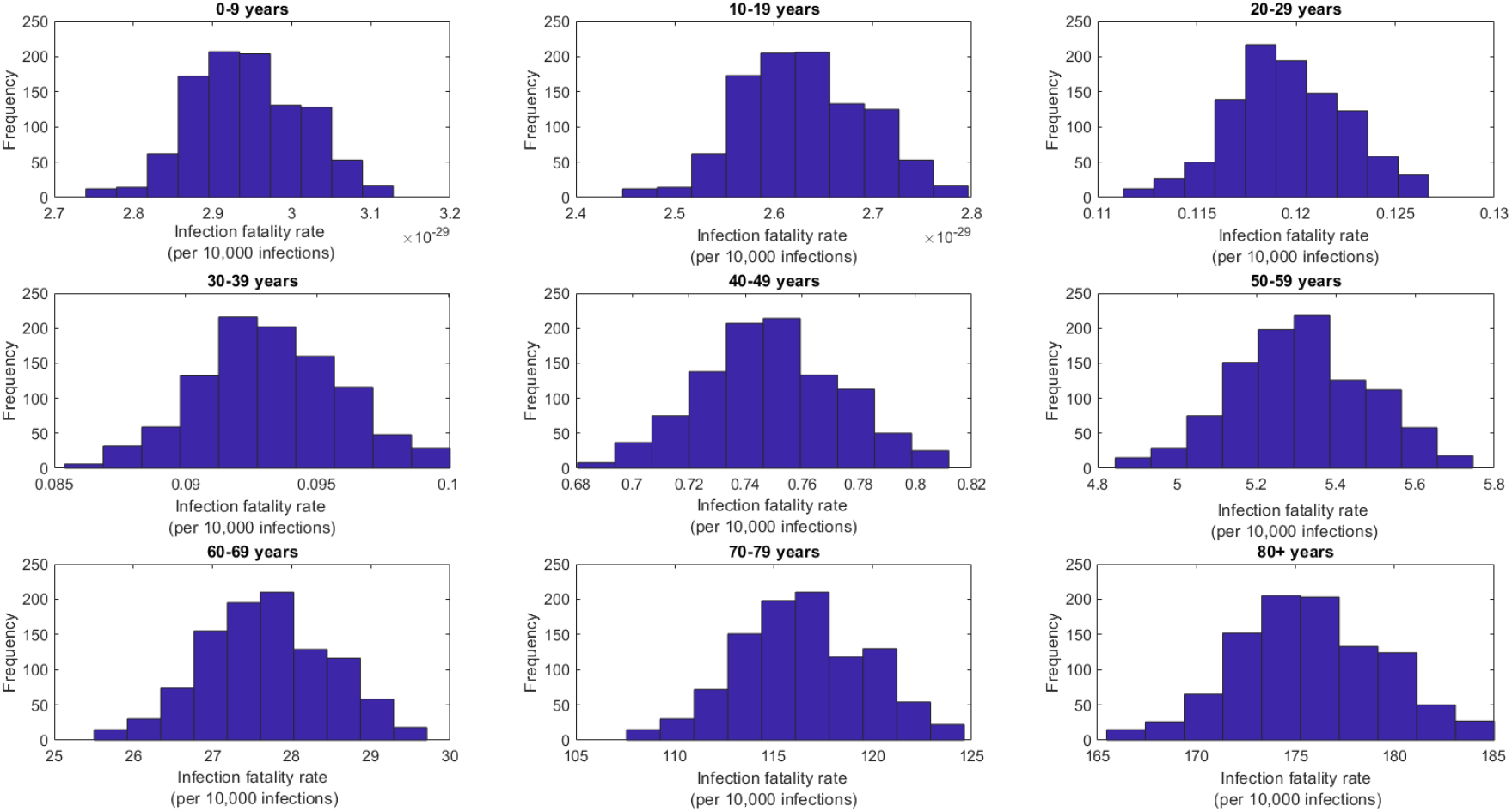
Posterior distribution of the age-specific infection fatality rate.

**Figure S8.**
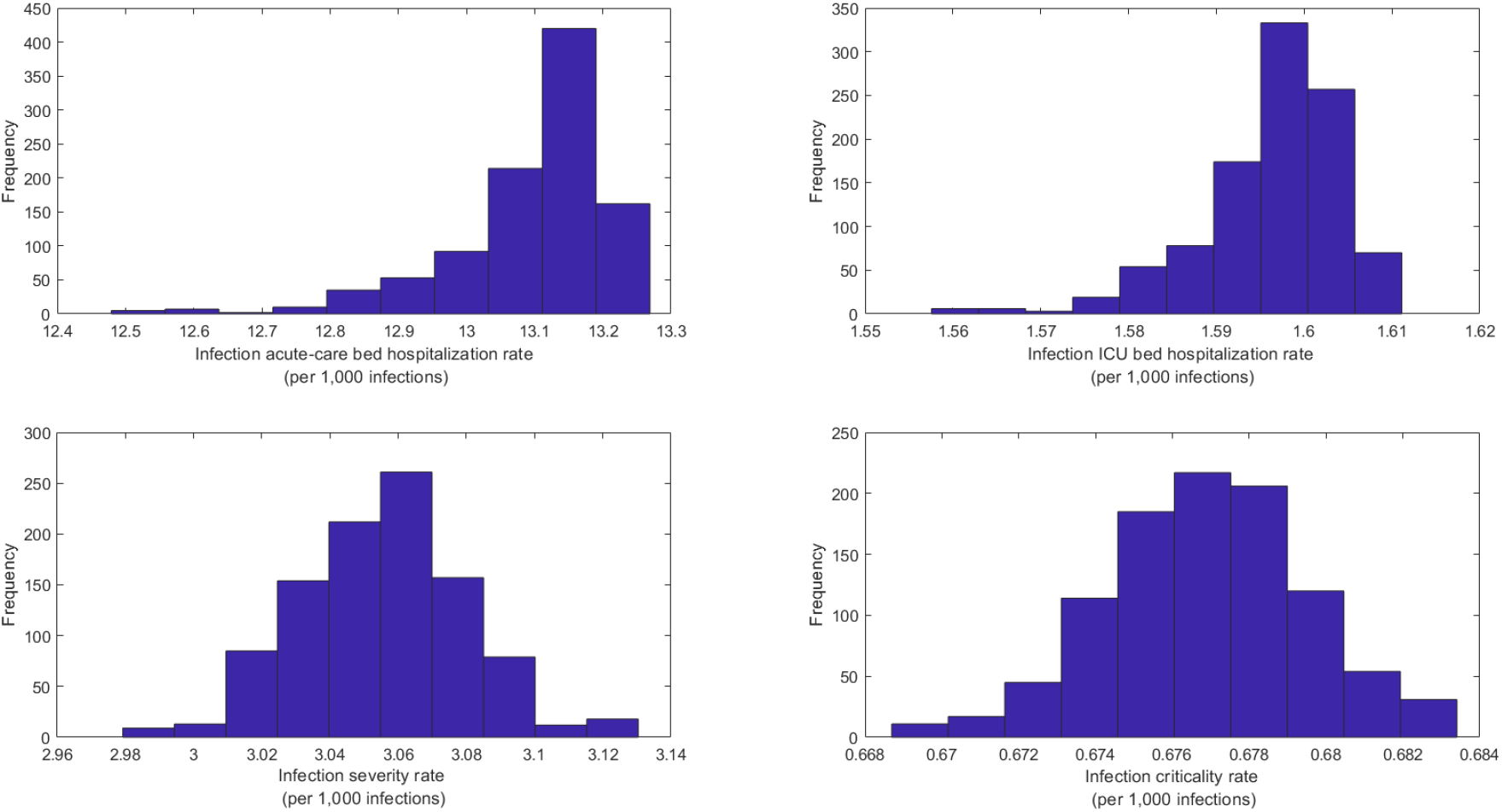
Posterior distribution of the overall (total population of all age groups) A) infection acute-care bed hospitalization rate, B) infection ICU bed hospitalization rate, C) infection severity rate, and D) infection criticality rate.

**Figure S9.**
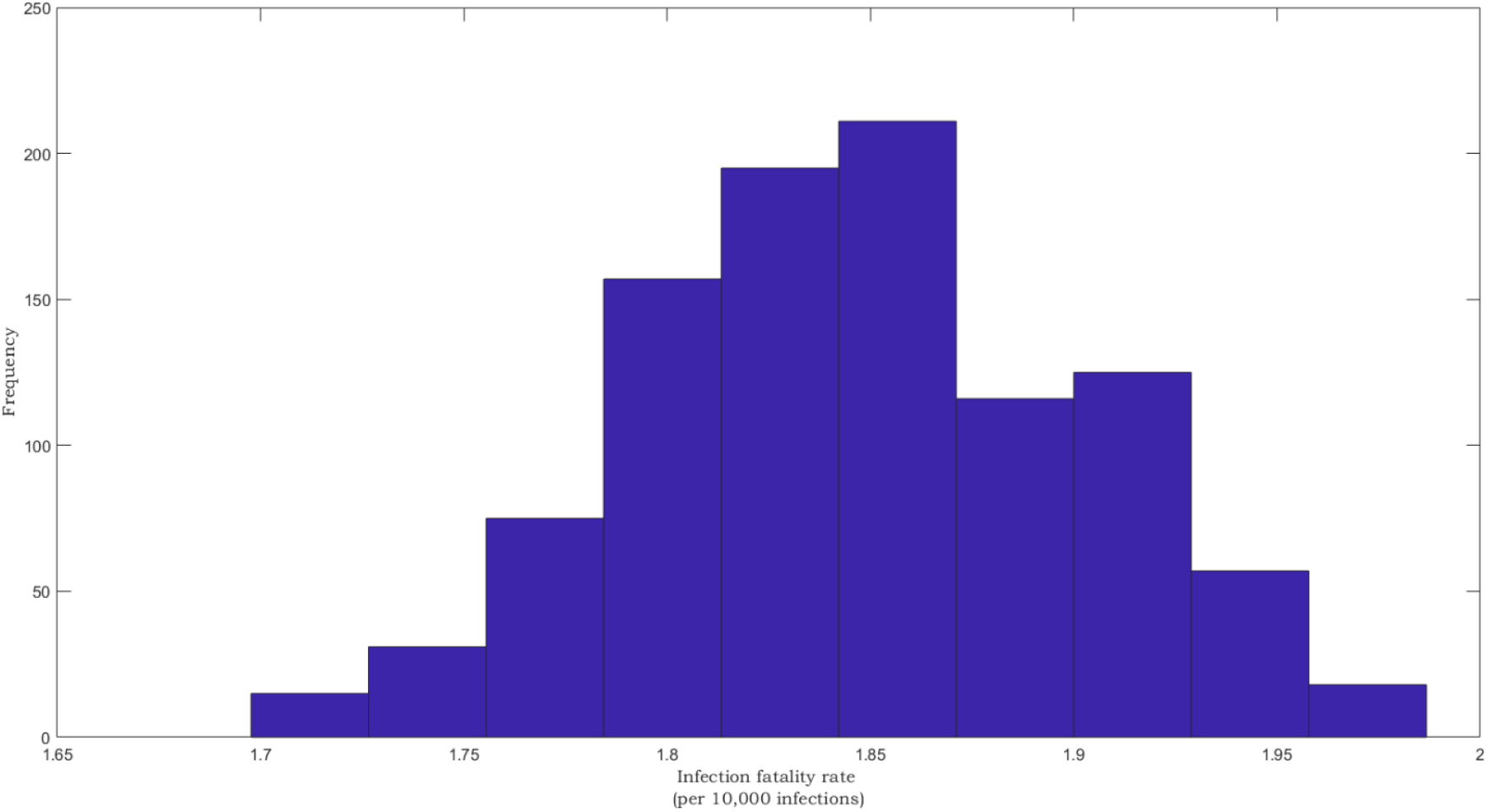
Posterior distribution of the overall (total population of all age groups) infection fatality rate.

**Figure S10.**
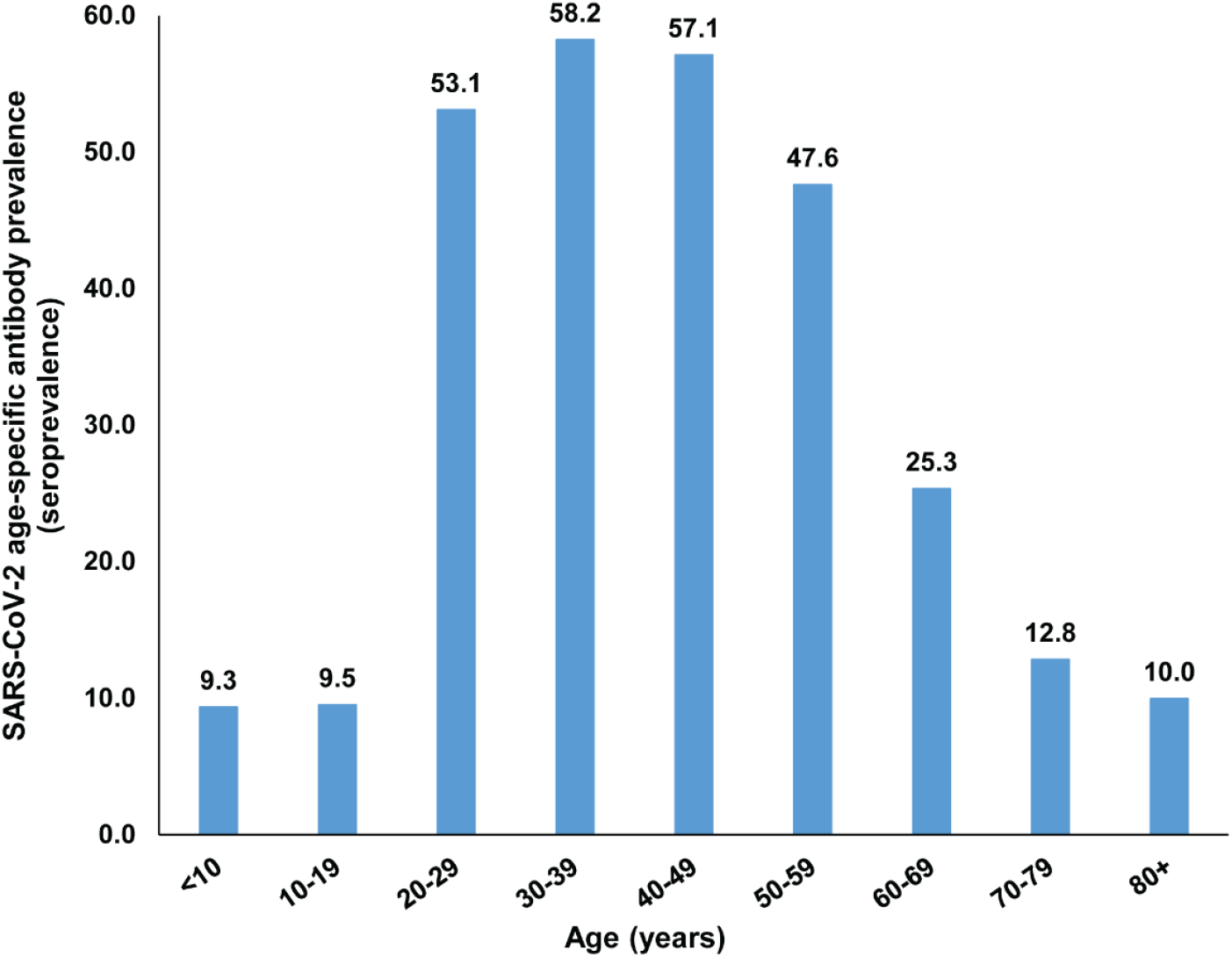
The SARS-CoV-2 age-specific antibody prevalence (seroprevalence) in the total population of Qatar based on compilation and analysis of seroprevalence data from a series of serological studies [3, 16-19].

## Notes

### Competing Interest Statement

The authors have declared no competing interest.

### Author Declarations

Studies were approved by Hamad Medical Corporation and Weill Cornell Medicine-Qatar Institutional Review Boards.

